# Cervical Microbiota Diversity and Functional Shifts in the Development of Cervical High-Grade Squamous Intraepithelial Lesions

**DOI:** 10.1101/2025.03.27.25324816

**Authors:** Marta Rosas Cancio-Suárez, Elena Moreno, Cristina del Valle Rubido, Marta Salvador, Ana Moreno, Laura Luna, Claudio Díaz-García, Carlos Tapia, Ana del Amo, Santiago Moreno, Matilde Sánchez-Conde, Sergio Serrano-Villar

## Abstract

Research on microbial changes in the cervix, where most human papillomavirus (HPV) complications arise, is limited. Here, we aimed to understand the specific role of the cervicovaginal microbiota in developing high-grade squamous intraepithelial lesions (HSIL) associated with HPV infection.

Our results show higher diversity in the microbiota associated with HSIL with the genus *Parvimonas, Fastidiosipila*, and *Pseudomonas* being the most abundant. Additionally, an imputed functional analysis revealed that pathways such as glycine, serine, threonine, and sulfur metabolism were enriched in cervical samples from women with HSIL. Identifying biomarkers that help prevent HSIL progression could benefit women at risk for developing HPV-related cancerous lesions.

## BACKGROUND

The cervicovaginal microbiota has co-evolved with mucosal immune cells to facilitate fetal implantation and development during pregnancy and protect the female urogenital tract from infection (1). The correlation between cervicovaginal microbiota and its role in either fostering susceptibility or providing protection against HPV, the third leading cause of cervical cancer, is gaining attention (2).

Research on bacterial communities has revealed that the optimal vaginal microbiota is predominantly composed of *Lactobacillus crispatus, L. gasseri*, and *L. jenseni*. These species secrete lactobiocins and biosurfactants that prevent other species from binding to the epithelium (3). Interestingly, a vaginal microbiota dominated by *L. iners* has been associated with higher odds of HPV prevalence, including high-risk HPV and cervical cancer or dysplasia (1,4). It has been established that the interaction between HPV, host immunity, and the vaginal microenvironment influences the risk of persistent HPV infection and the progression of cervical intraepithelial neoplasia (CIN) (5). However, while HPV-related complications primarily develop in the cervix, the cervical microbiota has been less studied.

Therefore, we conducted a study to explore the connections between HPV infection and squamous intraepithelial lesions, focusing on the composition and function of the cervical microbiota. We aimed to identify metabolites that can help identify risk markers for malignant or premalignant lesions, enhance current screening efforts, and identify potential therapeutic targets in the long term.

## METHODS

### Ethics

This study conformed to the principles of the Declaration of Helsinki and Good Clinical Practice Guidelines and was approved by the Independent Ethics Committee of the coordinating center Hospital Ramón y Cajal (, approval 146-21l) and the participating centers. All participants provided written informed consent before the initiation of the study procedures. For participants under 18 years of age, written informed consent was obtained from their parents or legal guardians.

### Study population

We conducted an observational, prospective, single-center study. The recruitment period for this study started on July 1, 2022 and ended on July 31, 2023. We systematically invited women undergoing routine cervical cytology at their primary care center or gynecology department assigned to the project in Madrid, Spain. We included 105 women classified into three groups: women who tested negative for HPV and had normal cytology (NEG), those who tested positive for HPV and had low-grade squamous intraepithelial lesions (LSIL), and those who tested positive and had high-grade squamous intraepithelial lesions (HSIL).

All the women provided written consent to participate in the study. All participants underwent endocervical sampling using Dacron swabs.

### 16S sequencing analysis

Cryopreserved samples obtained in sterile iSWAB-Microbiome tubes were thawed and DNA was extracted using the QIAamp DNA Mini Kit. For each sample, the V3-V4 regions of the 16S rRNA gene were analyzed using Illumina MiSeq (Novogene Bioinformatics Technology Co., Ltd.). 50 K reads with 250bp Pair-Ends inserts were analyzed for OTUs cluster and phylogenetic relationship construction. Different Alpha and Beta diversity indexes were calculated using Simpson, Shannon, and Principal Coordinate Analysis (PCoA) respectively. Differential abundance and function prediction analyses were performed by analyzing the compositions of microbiomes with bias correction (ANCOM-BC2) (6), identifying significantly different orthologous genes using the Kyoto Encyclopedia of Genes and Genomes (KEGG/KO) and Enzyme Commission (EC) terms between groups in the dataset. Bioinformatic analysis was performed using the Novogene pipeline. We then selected these terms and used MicrobiomeAnalyst to identify the enriched metabolic pathways or biological processes associated with them.

### Statistical analysis

Study data were collected and managed using REDCap electronic data capture tools v13.1.27. We report qualitative variables as frequency distribution and quantitative variables as medians with their 25^th^-75^th^ percentiles (P25-P75) or as means and standard deviation (SD), according to the data distribution. We performed comparisons between groups using the Chi-square test for categorical variables. Since the distribution of all the assessed variables deviated from normality, we used the Mann-Whitney U test for between-group comparisons of continuous variables. We used Stata v18.0 (StataCorp LP, College Station, TX, USA) to perform the statistical tests.

## RESULTS

### General characteristics

We included 105 women: 53, 34, and 18 in the NEG, LSIL, and HSIL groups, respectively. The median age of the participants was similar across the three groups: 42 years in the normal and HPV-negative cytology (NEG) group, 41 years in the LSIL group, and 38.5 years in the HSIL group. Most participants were Spanish (94%, 74 %, and 72% of the NEG, LSIL, and HSIL groups, respectively). A smaller proportion of participants were from Eastern Europe and South America.

We found differences among the groups in terms of health determinants such as smoking and HPV vaccination. Women in the LSIL and HSIL groups had a higher prevalence of current smoking than those in the NEG group did. Additionally, there were notable differences in HPV vaccination rates, with a higher percentage of vaccinated women in the NEG and LSIL groups than that in the HSIL group.

Among HPV-seropositive women, the distribution of HPV types was similar between the LSIL and HSIL groups (p = 0.16). Most high-risk HPV genotypes were neither 16 nor 18. For these high-risk HPV genotypes, the specific type was not identified by polymerase chain reaction (PCR), which means that it could be any of the following: 31, 33, 35, 39, 45, 45, 51, 52, 56, 58, 59, 59, 66, and 68. For more information, please refer **to Table S1**.

### Microbiota diversity and specific taxa associations in the development of cervical HSIL

To understand the influence of microbiota composition on the development of CC in the context of HPV infection, we performed microbiota analyses by dividing the samples into two groups: women who developed HSIL (YesHSIL) and those who did not develop HSIL (NoHSIL). Focusing on extreme phenotypes may help identify more robust microbial markers.

To measure the richness and evenness of bacterial taxa within a community, we calculated alpha diversity indexes (**Figures 1A** and **B)**. Alpha diversity, calculated using Shannon and Simpson indexes, varied significantly between the YesHSIL and noHSIL groups. We also applied a dimensionality-reduction technique, Principal Coordinates Analysis (PCoA), to visualize the differences between microbial community configurations (beta diversity), and we found no clearly distinct clusters (**Figure 1C**). To identify specific taxa that differed significantly in abundance between groups, we used the ANCOM-BC2 method to compare the differences in absolute abundances. Women with HSIL showed increased abundances in the genus *Parvimonas, Fastidiosipila*, and *Pseudomonas*, while the genera *Porphyromonas, Elizabethkingia, Bacteroides*, and *Akkermansia* were depleted (**Figure 1D**).

**Figure 1:**
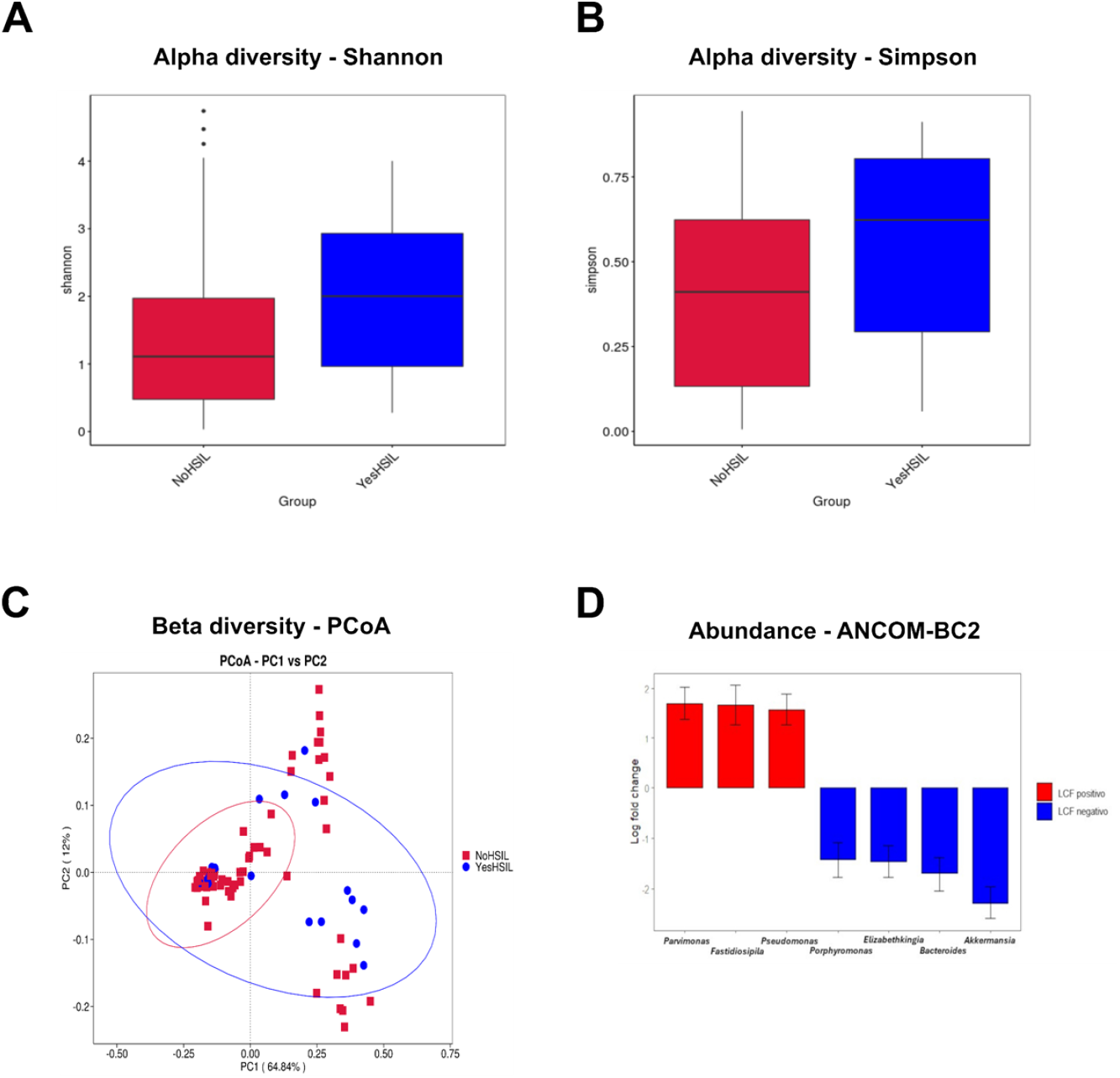
Diversity and abundance of cervicovaginal microbiota in the two compared groups (NoHSIL and YesHSIL). **A**. Boxplots based on Shannon index, showing the maximum, minimum, median and abnormal values of the index from each group. **B**. Boxplots based on Simpson index, showing the maximum, minimum, median and abnormal values of the index from each group. Significant differences are shown with a line and asterisk (p = 0.02, Kruskal-Wallis test). **C**. PCoA based on the weighted Unifrac distance showing the two components explaining most of the variance (PC1 and PC2). The multiresponse permutation procedure (MRPP) test was performed to statistically evaluate the differences between the groups (MRPP significance: NoHSIL-YesHSIL = 0.238). **D**. The ANCOM-BC2 test was performed to analyze the abundance of genera in the two groups. The results were calculated as Log Fold Change (LFC). Red indicates genera with a positive LFC (higher abundance) in the HSIL group. Blue indicates genera with a negative LFC (lower abundance) in the HSIL group.

### Functional analysis of the cervical microbiome reveals key pathways associated with the development of cervical HSIL

Next, we sought to understand the potential mechanisms by which the cervical microbiome influences the development of HSIL. We compared the bacterial imputed functions between the most extreme phenotypes, the NEG and YesHSIL groups. We also applied the ANCOM-BC2 method, focusing on the KEGG and EC databases, to identify the most significant terms. We identified 37 KO and 17 EC terms that were significantly associated with HSIL (**Figure 2** and **Table S2**).

**Figure 2:**
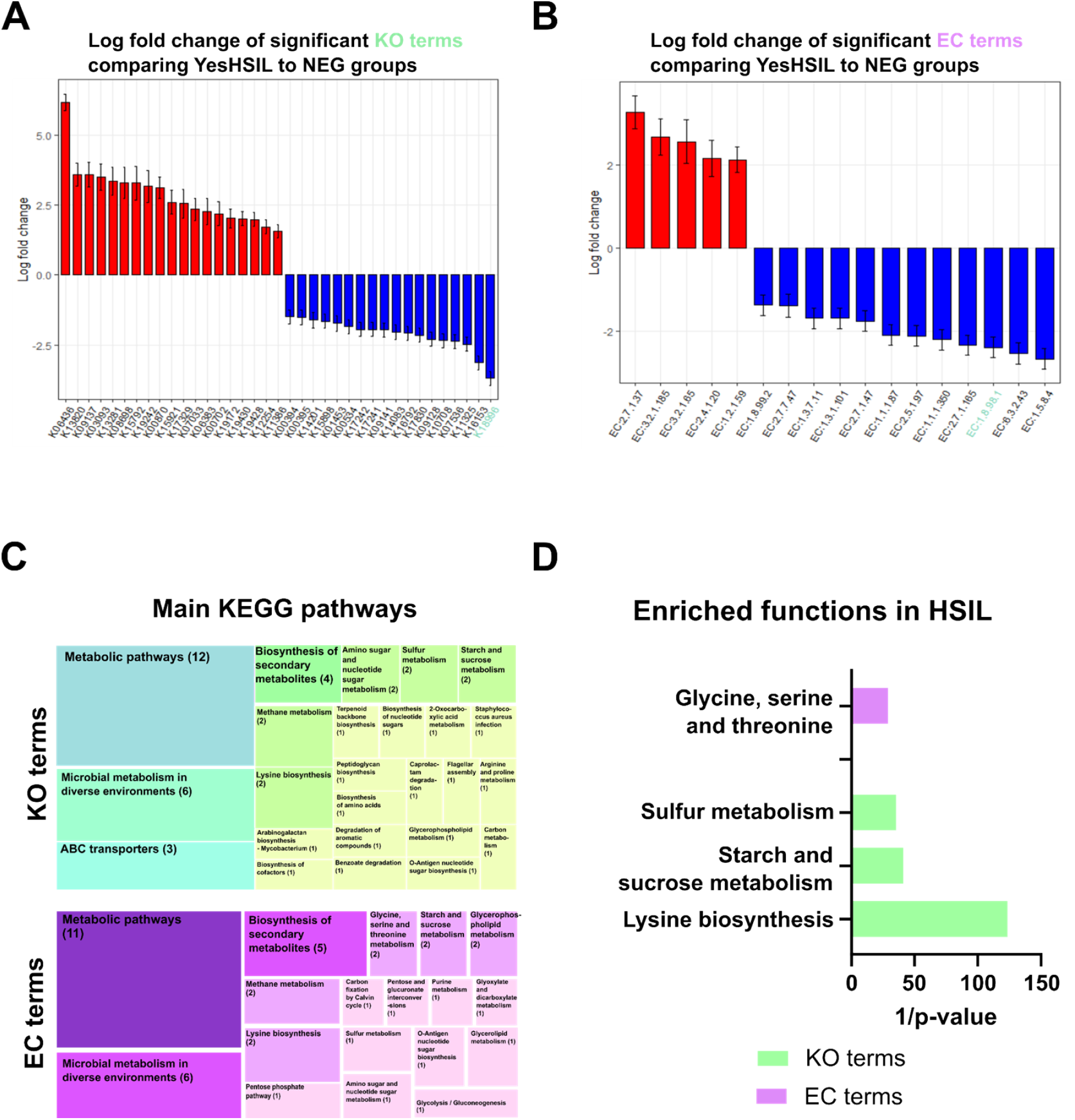
Functional terms differentially found in the cervical HSIL group compared to the negative group. **A**. KEGG terms obtained using ANCOM-BC2 and selected based on their LFC. **B**. EC terms obtained using ANCOM-BC2 and selected based on their LFC. The genus in red had a positive LFC (higher abundance) in the group with HSIL. The genus in blue had a negative LFC (lower abundance) in the HSIL group. Genus subjected to pseudocount correction by ANCOM-BC2 is shown in green. **C**. The main terms were obtained using KEGG mapper with the list of significant terms from the ANCOM-BC2 analysis. Numbers in parentheses denote the number of terms in each category. **D**. Main terms obtained after performing enrichment analysis, as explained in the Methods section for the KO terms (green) and EC terms (purple) obtained using ANCOM-BC2. The inverse of the p-value is represented on the x-axis for visualization purposes.

This two-step approach allows us to first identify relevant genes through differential abundance Analysis and interpretation of the functional impact of these genes were performed by mapping them to pathways, reducing noise, and increasing the biological relevance of our results. In women with HSIL, several KO terms related to metabolic pathways, biosynthesis of secondary metabolites, and microbial metabolism were significantly enriched, including those associated with amino sugar and nucleotide metabolism, sulfur metabolism, and starch/sucrose metabolism. On the enzyme side, key EC terms related to lysine biosynthesis, carbohydrate degradation, and glyoxylate metabolism were more abundant, whereas pathways such as glycine/serine/threonine metabolism were depleted.

## DISCUSSION

Our study revealed a more diverse vaginal microbiota in women with cervical dysplasia than that in healthy women, with differences in specific taxa and biological functions linked to the development of cervical HSIL. The increased abundance of *Parvimonas, Fastidiosipila*, and *Pseudomonas* in women with HSIL suggests their potential role in cervical dysbiosis and HPV progression. Notably, *Parvimonas spp*. has been frequently associated with precancerous lesions and dysbiosis (7,8), particularly in connection with sexually transmitted infections such as *Chlamydia trachomatis* and persistent HPV infection. Although *Fastidiosipila* has been less studied, our findings align with previous research linking it to HSIL and CIN (9). Interestingly, *Pseudomonas*, while often considered part of the healthy endocervical microbiota, has been linked to various pathologies such as *Chlamydia trachomatis* infection (10,11), further supporting the idea that shifts in the microbiota may contribute to disease. The prominence of these genera, which are more commonly associated with the intestinal microbiota, suggests possible translocation or shared pathways between the gut and the cervicovaginal environment (12). Mechanisms such as bacterial ascent through the reproductive tract or hematogenous spread from gastrointestinal niches could explain this crossover. However, further studies are required to explore these possibilities (13).

The functional analysis further reinforces the association between microbial dysbiosis and HSIL. We found enriched gene pathways related to typical bacterial metabolic processes and enzyme pathways related to glycine, serine, and threonine metabolism, suggesting that the microbial community in HSIL cases exhibits heightened metabolic activity. The increase in glycosidases and a corresponding decrease in oxidoreductases also point to significant metabolic alterations, potentially contributing to a dysbiotic environment conducive to lesion progression. These findings are consistent with metabolic dysregulation linked to cancer development, including processes such as the Warburg effect and oxidative stress, which may promote DNA damage and promote the development of cancerous lesions (14,15).

However, our study has some limitations. However, our study has some limitations. Although we controlled for variables such as menopausal status and sex hormones, other factors such as oral contraceptive use and family history of cancer were not accounted for, which may have influenced changes in the cervical microbiota. Additionally, most studies examining the relationship between microbiota and cervical dysplasia have focused on vaginal rather than cervical samples (3). Future studies correlating both environments could provide a less invasive means of assessing women at a higher risk of progressing to HSIL by linking findings in the vaginal microbiota to those in the cervix.

Geographic location may also influence microbiota composition (16). To the best of our knowledge, this study is the first to examine the cervical microbiota in women from Spain, and direct comparisons with the existing literature are challenging. This underscores the need for region-specific studies to better understand the microbial landscape and its relationship to cervical dysplasia. Lastly, although our functional analysis identified significant pathways, the presence of genes does not guarantee their activity, highlighting the importance of further experimental studies, particularly longitudinal studies, to establish causal relationships between cervical dysbiosis and CIN progression.

Despite these limitations, our study lays the groundwork for future research to explore the role of the cervicovaginal microbiome in HSIL development. The microbial taxa and metabolic pathways identified here could inform studies focused on identifying markers for cervical dysplasia progression, offering avenues for targeted screening and therapeutic interventions.

## Acknowledgments

We thank all the patients and healthcare workers who participated in the study.

## Author’s contributions

Conceptualizations: M.R.C-S, S.S-V; Methodology: M.R.C-S; L.L., E.M., M.F., A.A.; Patient recruitment: M.R.C-S, L.L., MS-C, C.V.R; Laboratory measurements: E.M. M.F., A.A.; Interpretation of bioinformatic analysis: E.M; Project supervision: M.S-C, S.S-V.; Writing – original draft: M.R.C-S, E.M.; Writing – review and editing: all authors

## Data Availability

The datasets generated and/or analysed during the current study are available in the European Nucleotide Archive (ENA) at EMBL-EBI under accession number PRJEB83336/ERP166979 (https://www.ebi.ac.uk/ena/browser/view/PRJEB83336). Datasets will be make public once publication is accepted.

